# Preferred left ventricular lead position for upgrade from right ventricular pacing to cardiac resynchronization therapy

**DOI:** 10.1101/2023.04.27.23289232

**Authors:** Michio Ogano, Yu-ki Iwasaki, Taiji Okada, Jun Tanabe, Wataru Shimizu, Kuniya Asai

**Affiliations:** Department of Cardiovascular Medicine, Shizuoka Medical Center, 762-1 Nagasawa, Shimizu, Sunto Shizuoka 4110906, Japan; Department of Cardiovascular Medicine, Nippon Medical School, 1-1-5 Sendagi, Bunkyo Tokyo 1138603, Japan

**Keywords:** cardiac resynchronization therapy, cardiac resynchronization therapy upgrade, left ventricle lead position, left ventricular electrical delay, acute hemodynamic response

## Abstract

**Background:** Cardiac resynchronization therapy (CRT) is a well-established treatment for symptomatic heart failure with electrical dyssynchrony. The left ventricle (LV) lead position is recommended at LV posterolateral to lateral sites in patients with left bundle branch block; however, its preferred region remains unclear in patients upgrading from conventional right ventricular (RV) apical pacing to CRT. We aimed to identify the preferred LV lead position for upgrading conventional RV apical pacing to CRT.

**Methods:** This study used electrode catheters positioned at the RV apex and LV anterolateral and posterolateral sites via the coronary sinus (CS) branches to measure the ratio of activation time to QRS duration from the RV apex to the LV anterolateral and LV posterolateral sites during RV apical pacing. We performed simultaneous biventricular pacing at the RV apex and each LV site and measured the differences in QRS duration and LV dP/dt_max_ from those of RV apical pacing.

**Results:** This study included 37 patients with anterolateral and posterolateral LV CS branches. During RV apical pacing, the average ratio of activation time to QRS duration was higher at the LV anterolateral site than at the LV posterolateral site (0.90±0.06 vs. 0.71±0.11, p<0.001). The decreasing ratio of QRS duration and the increasing ratio of LV dP/dt_max_ were higher at the LV anterolateral site than at the LV posterolateral site (45.7±18.0% vs. 32.0±17.6%, p<0.001 and 12.7±2.9% vs. 3.7±8.2%, p<0.001, respectively) during biventricular pacing compared with those during RV apical pacing.

**Conclusions:** The LV lead position is preferred at the LV anterolateral site in patients upgrading from conventional RV apical pacing to CRT.

**Condensed abstract:** Pacing at the latest electrical activation site is crucial to improve electrical dyssynchrony in cardiac resynchronization therapy (CRT). However, the preferred location of the left ventricle (LV) lead position in patients upgrading from conventional right ventricular apical pacing to CRT is unclear. This study aimed to investigate proper strategies for CRT and identify an approach for patients upgrading to CRT. We showed that the preferred location is the LV anterolateral site. Our findings will help cardiologists and clinicians develop better strategies for treating patients with heart failure complicated with atrioventricular block.

## Introduction

Cardiac resynchronization therapy (CRT) is a well-established treatment for patients with symptomatic heart failure (HF) and electrical dyssynchrony. In previous large clinical trials with CRT, the left ventricle (LV) lead position was instructed to be at the posterolateral to lateral branch of the coronary sinus (CS) (1,2). Therefore, the position from the posterolateral to the lateral branch of the CS is presumably ideal for LV lead placement. However, there were differences in the underlying electrical dyssynchrony in individual patients (3), and the optimal LV pacing site should be individually determined.

Several studies showed that a predictor of symptomatic and structural response to CRT is the most electrically delayed site in the LV that is assessed by the electrical delay from the beginning of the native QRS complex to the local LV electrogram (Q-LV) or by the Q-LV ratio, defined as Q-LV over the QRS duration (QRSd) (4,5). This finding is consistent with the concept that CRT eliminates electrical dyssynchrony. Right ventricular (RV) apical pacing leads to nonphysiological electrical myocardial conduction and mechanical left ventricular dyssynchrony (6). Some patients may have LV systolic dysfunction, resulting in HF (7,8), and they are recommended to upgrade to CRT by adding a new LV lead (9,10). However, the electrically optimal LV lead site remains unclear in patients planned to upgrade from conventional RV apical pacing to CRT.

Thus, we aimed to identify the preferred LV lead position for upgrading conventional RV apical pacing to CRT.

## Methods

### Patients

This study included consecutive patients older than 65 years who were scheduled to undergo catheter ablation for atrial fibrillation between March 2022 and December 2022. All patients underwent a 12-lead electrocardiogram (ECG,) laboratory tests, and echocardiography. The exclusion criteria were as follows: 1) bundle branch block, 2) post-pacemaker implantation state, 3) history of myocardial infarction or surgery for cardiovascular diseases, and 4) severe renal function damage, defined as an estimated glomerular filtration rate of <30 mL/min/1.73 m^2^.

This study was approved by the Center Ethics Committee in our hospital (2021-R31), conformed to the ethical guidelines of the 1975 Declaration of Helsinki, and was approved by the institution’s human research committee. Written informed consent was obtained from all participants.

### Electrophysiological study and acute hemodynamic measurements with temporary pacing

Antiarrhythmic drugs were discontinued for at least five half-lives before catheter ablation. Pulmonary vein isolation was performed in all the patients using radiofrequency currents or cryoballoons. Additional superior vena cava isolation or cavotricuspid isthmus ablation was performed at the discretion of the operator. If sinus rhythm could not be obtained after pulmonary vein isolation, intracardiac defibrillation was performed to restore sinus rhythm.

After catheter ablation for atrial fibrillation, electrophysiological studies and acute hemodynamic measurements were performed with temporary pacing. Angiography of the CS using a balloon-occlusion catheter (Wedge pressure catheter, Telefrex, PA, USA) to delineate the CS branches was eligible for LV lead deployment. A pigtail-shaped catheter (Heartcass; Terumo, Tokyo, Japan) was introduced into the LV via a transseptal approach for hemodynamic measurements. The electrode catheter was placed in the high atrium and the RV apex. The CS was cannulated with an 8.5-Fr pre-shaped SL2 sheath (Abbott Medical, St Paul, MN, USA). A 3.5-Fr over-the-wire-type pacing catheter (InterNova Monorail Catheter; InterNova, Tokyo, Japan) with a 0.014-inch guidewire was placed in each CS branch.

The baseline maximum LV pressure rate (LV-dP/dt_max_) was measured during pacing in the DDI mode performed from the RV apex. Atrial pacing was programmed at 10 beats/min above the intrinsic rate. The atrioventricular (AV) delay was set at 100 ms below the baseline AV delay to ensure consistent ventricular capture. The electrical delay to each CS branch during RV pacing was measured as the Q-LV interval, and the ratio of Q-LV to QRSd was calculated as the QRS ratio. Simultaneous RV pacing and LV pacing were continuously performed, and changes in QRSd and LV dP/dt_max_ were measured. LV dP/dt_max_ was calculated electrically from every heartbeat for at least 10 s under steady-state conditions.

### Location of the CS branches

The CS branches were identified based on CS angiography. CS angiography was performed in two orthogonal views (right anterior oblique, 30°; left anterior oblique, 45°). In the right anterior oblique view, which represented the longitudinal axis of the heart, the areas were classified into three segments: basal, midventricular, and apical. In the left anterior oblique view, which represented the short axis of the heart, a line between the anterior interventricular vein and the middle cardiac vein was drawn. From the anterior interventricular line, 0°-30°, 30°-90°, 90°-150°, and 150°-180° were defined as the anterior, anterolateral, posterolateral, and posterior areas, respectively (Figure 1).

**Figure 1.**
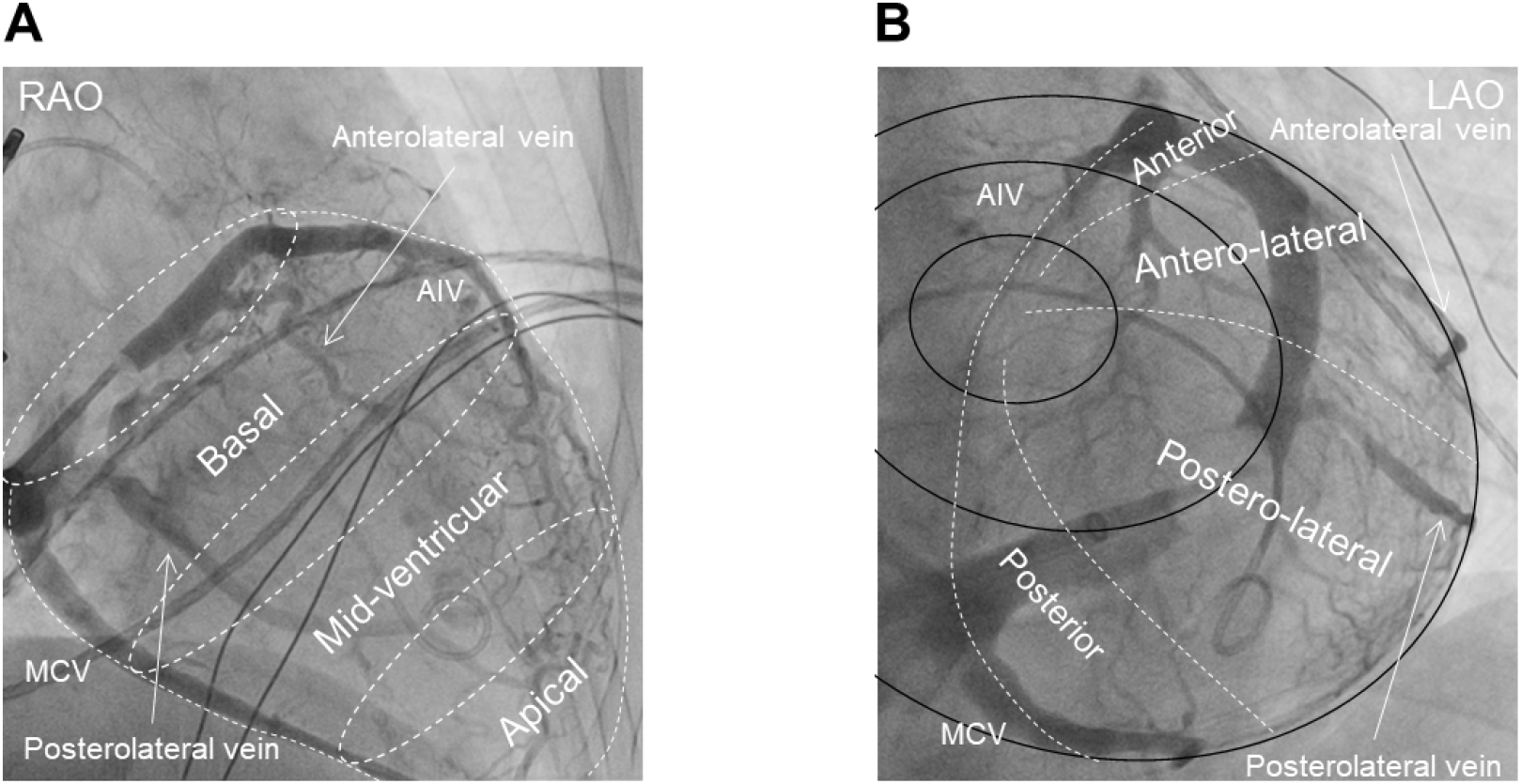
Coronary sinus venography to classify possible left ventricular lead positions. **(A)** Right anterior oblique (RAO) view representative of the long axis of the heart. These areas are divided into three segments: basal, midventricular, and apical. **(B)** Left anterior oblique (LAO) view representative of the short axis of the heart. After drawing a line between the anterior interventricular vein (AIV) and the middle cardiac vein (MCV), the areas are divided into four segments based on the angle from the center: anterior (0°–30°), anterolateral (30°–90°), posterolateral (90°–150°), and posterior (150°–180°).

### Statistical analyses

Continuous data were expressed as the mean and standard deviation and were compared using the Mann–Whitney U test or Wilcoxon signed-rank test, as appropriate. Meanwhile, categorical variables were expressed as absolute numbers and percentages and were compared using Fisher’s exact test. Assuming a two-sided alpha of 0.05, a statistical power of 80%, and an effect size of 0.5, the target sample size was calculated to be 34. To account for a potential dropout rate of 5%, we calculated that the trial would need to include 37 patients. All statistical analyses were performed using the Statistical Package for Social Science (version 24.0; IBM, Armonk, NY, USA) and R statistical software version 4.0.2 (R Foundation for Statistical Computing, Vienna, Austria). Statistical significance was defined as a two-tailed p<0.05.

## Results

### Patient background

Among the 113 patients enrolled to undergo catheter ablation for atrial fibrillation, 47 patients were excluded from this study because of age ≤64 years (n=32), history of myocardial infarction or surgery for cardiovascular diseases (n=8), bundle branch block (n=5), and post-pacemaker implantation state (n=2). The remaining 56 patients underwent CS angiography, among whom 37 patients (66.1%) showed anterolateral and posterolateral branches in the CS and were eligible for LV lead implantation (Figure 2). The patient characteristics are presented in Table 1.

**Figure 2.**
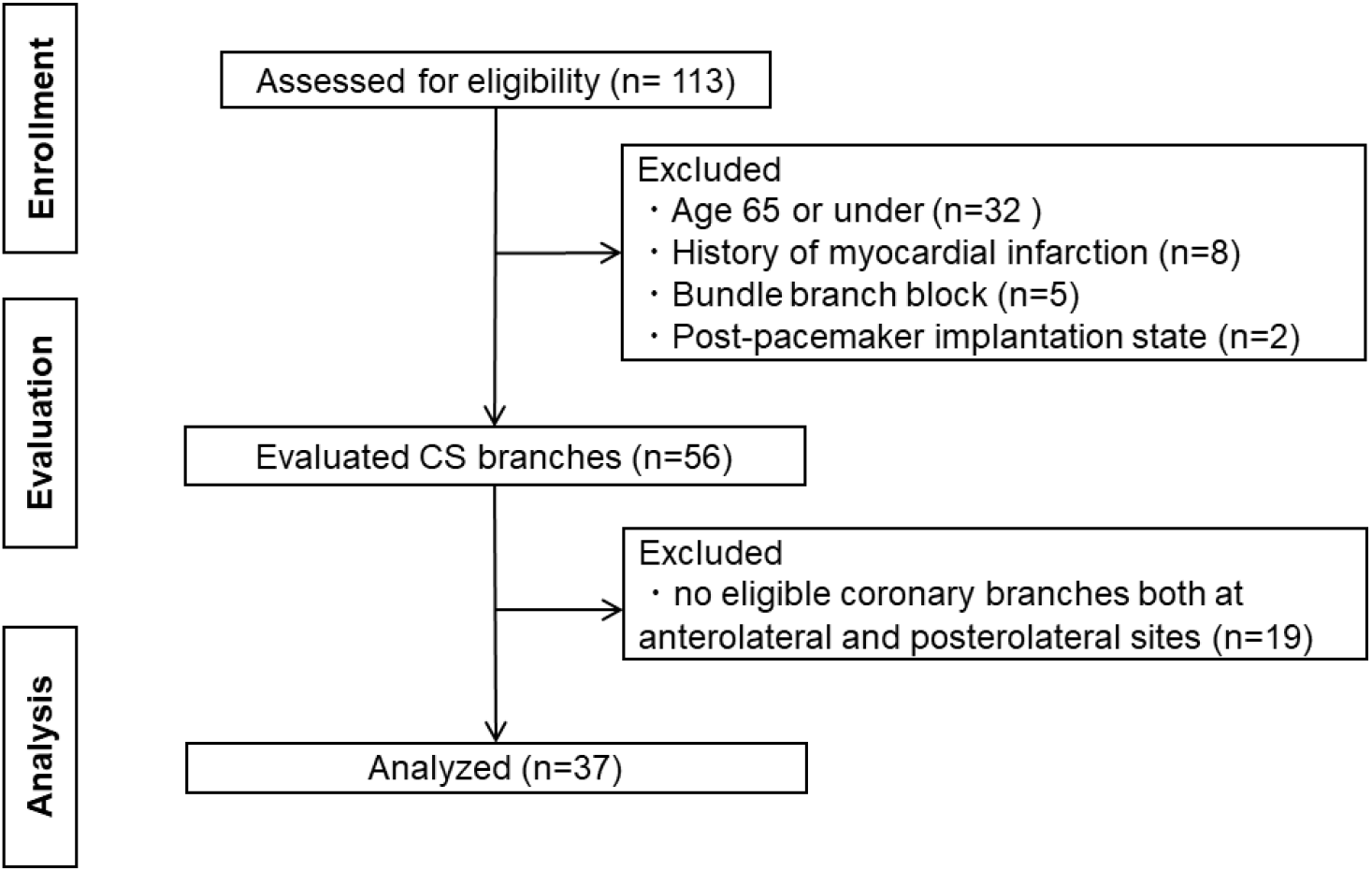
Patient selection flow chart. CS: coronary sinus

**Table 1.** Baseline patient characteristics.

| Characteristics | Total<br>(N = 37) |
| --- | --- |
| Male sex | 26 (70.3) |
| Age (years) | 75 ± 8 |
| Intrinsic QRS (ms) | 97 ± 6 |
| Intrinsic HR (bpm) | 65 ± 12 |
| Paroxysmal atrial fibrillation | 20 (54.1) |
| Hypertension | 29 (78.4) |
| Diabetes mellitus | 12 (32.4) |
| Dyslipidemia | 16 (43.2) |
| Heart failure | 13 (35.1) |
| LVEF (%) | 58.3 ± 9.3 |
| LVEDV (ml) | 95.7 ± 23.8 |
| LVESV (ml) | 40.7 ± 17.1 |
| IVST (mm) | 9.3 ± 1.4 |
| PWT (mm) | 8.9 ± 1.0 |
| LAD (mm) | 38.9 ± 4.7 |
| E/e' | 12.1 ± 4.8 |
| BNP (pg/mL) | 133.4 ± 144.5 |
| β blocker use | 18 (48.6) |
| ACEI/ARB/ARNI use | 18 (48.6) |
| MRA use | 4 (10.8) |
| SGLT2I use | 6 (16.2) |

Data are presented as n (%) or as the mean ± SD.
HR, heart rate; LVEF, left ventricular ejection fraction; LVEDV, left ventricular end-diastolic volume; LVESV, left ventricular end-systolic volume; IVST, interventricular septum thickness; PWT, posterior left ventricular wall thickness; LAD, left atrial dimension; BNP, brain natriuretic peptide; ACEI, angiotensin-converting-enzyme inhibitor; ARB, angiotensin-receptor blocker; ARNI, angiotensin receptor neprilysin inhibitor; MRA, mineralocorticoid receptor antagonist; SGLT2I, sodium-glucose cotransporter 2 inhibitor

### Electrophysiological study and acute hemodynamic measurements with temporary pacing

All 37 patients showed regular sinus rhythm after catheter ablation for atrial fibrillation. The mean baseline heart rate was 65±12 beats/min. During pacing in the AAI mode (10 beats/min above the intrinsic rate), the average QRSd and LV dP/dt_max_ were 97±6 ms and 1192.2±312.5 mmHg/s, respectively. QRSd was significantly longer (97±6 ms vs. 159±13 ms, p<0.01) while LV dP/dt_max_ was significantly shorter (1192.2±312.5 mmHg/s vs. 1071.0±293.5 mmHg/s, p<0.01) during pacing in the DDI mode from the RV apical site, compared with those in the AAI mode.

The Q-LV interval as electrical delay to each CS branch during RV pacing was evaluated at the LV anterolateral and posterolateral branches via CS branches. First, we assessed the longest Q-LV interval site from the perspective of the LV longitudinal view: the right anterior oblique view. At the LV anterolateral branch, 36 (97.3%) and 1 (2.7%) patient showed the longest Q-LV interval site at the base and middle ventricles, respectively. At the LV posterolateral CS branch, 31 (83.8%) and 6 (16.2%) patients showed the longest Q-LV interval site at the base and middle ventricles, respectively. Notably, no patient showed the longest Q-LV interval site at the apex.

Then, we evaluated the Q-LV interval in the short-axis and left anterior oblique view. The Q-LV interval and the QLV ratio at the latest activation site were longer and higher at the LV anterolateral site than at the LV posterolateral site (144.2±14.4 ms vs. 114.3±18.3 ms, p<0.01 and 0.91±0.06 vs 0.72±0.711, p<0.01, respectively). LV stimulation was attempted at the longest Q-LV interval. However, the pacing threshold was too high to stimulate the LV in 9 (24.3%) and 7 (18.9%) patients at the LV anterolateral and posterolateral branches, respectively (Table 2). In these 16 patients, LV myocardial capture was performed after the pacing site was moved to a more apical site. There was no significant difference in the obtained Q-LV time from the latest activation site to the LV myocardial capture site between the LV anterolateral and posterolateral sites (7.3±3.6 ms and 7.9±6.1 ms, p=0.85). The Q-LV interval and QLV ratio at the pacing LV myocardial capture site were longer and higher at the LV anterolateral site than at the LV posterolateral site (142.5±15.2 ms vs. 112.6±8.7 ms, p<0.01 and 0.90±0.06 vs. 0.71±0.11, p<0.01, respectively).

**Table 2.** Difference between the latest activation site and the actual available pacing site.

|  | Anterolateral branch |  | Posterolateral branch |  |
| --- | --- | --- | --- | --- |
|  | Base | Mid-ventricle | Base | Mid-ventricle |
| Latest activation site | 36 | 1 | 34 | 3 |
| Actual pacing available site | 31 | 6 | 28 | 9 |

Simultaneous RV apex and LV pacing was performed at each LV anterolateral or LV posterolateral site. The QRSd was shorter at the LV anterolateral site than at the LV posterolateral site (114±11 ms vs. 127±12 ms, p<0.01). The decreasing ratio of QRSd in biventricular pacing compared with that in RV apical pacing was greater at the LV anterolateral site than at the LV posterolateral site (45.7±18.0% vs. 32.0±17.6%, p<0.001). The LV dP/dt_max_ was higher at the LV anterolateral site than at the LV posterolateral site (1188.8±291.3 mmHg/s vs. 1092.9±261.2 mmHg/s, p<0.01). The increasing ratio of LV dP/dt_max_ from the RV apical to biventricular pacing was also higher at the LV anterolateral site than at the LV posterolateral site (12.7±8.9% vs. 3.7±8.2%, p<0.01). A representative case is shown in Figure 3A-B.

**Figure 3.**
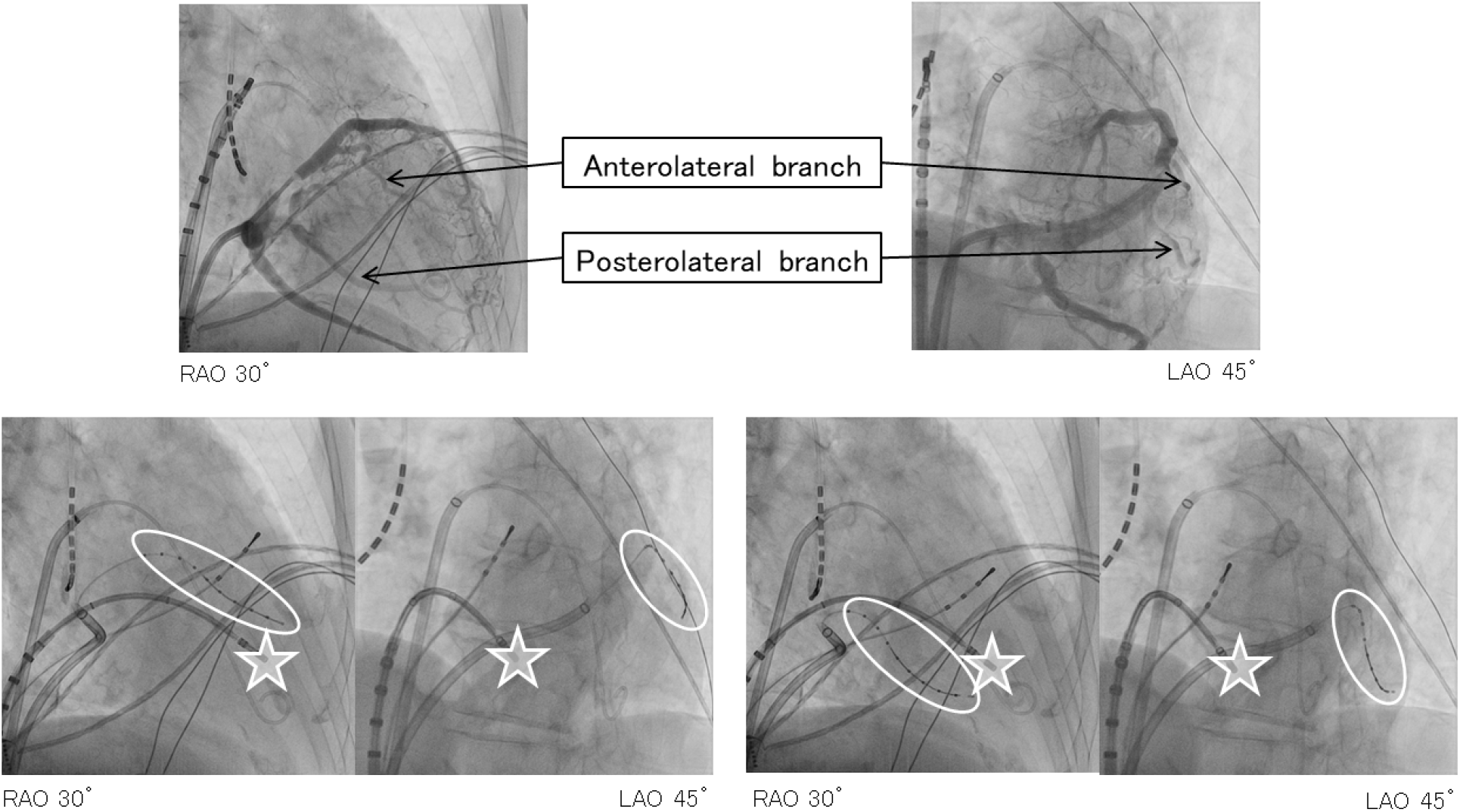
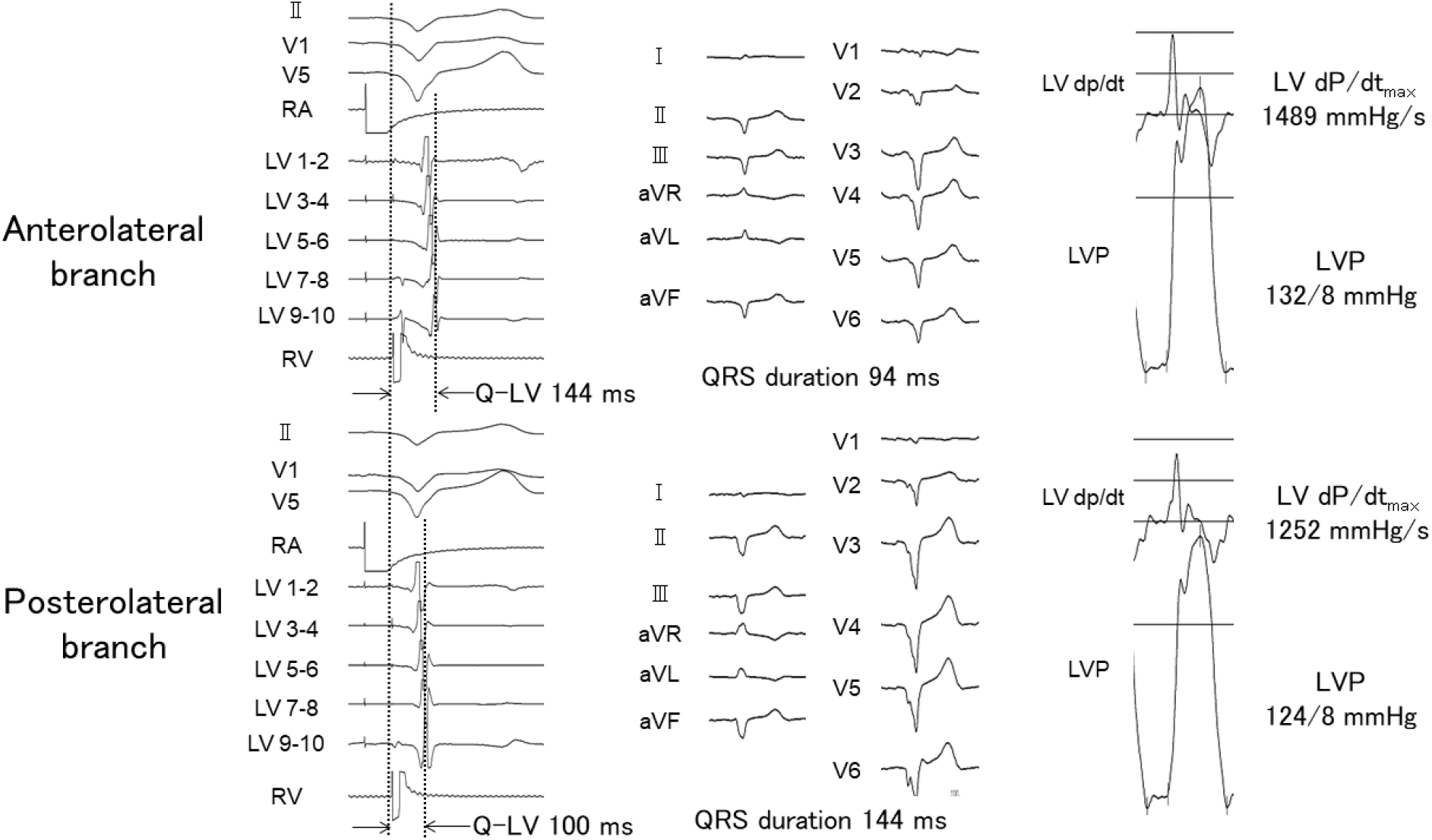
Example of electrophysiological studies and acute hemodynamic measurements. **(A)** Coronary sinus angiography is performed in an 80-year-old male. The anterolateral and posterolateral branches are depicted (upper panel). An electrode catheter is inserted into each branch (white circle). During right ventricular (RV) apical stimulation (white star), the Q-LV is measured at each branch (lower panel). **(B)** The Q-LV interval is longer at the left ventricular (LV) anterolateral site (144 ms) than at the LV posterolateral site (100 ms) (left panel). Simultaneous biventricular pacing shows that the QRS duration us shorter at the LV anterolateral site (94 ms) than at the LV posterolateral site (144 ms) (middle panel). The LV dP/dt_max_ and left ventricle pressure (LVP) are higher at the LV anterolateral site (1489 mmHg/s and 132/8mmHg) than at the LV posterolateral site (1252 mmHg/s and 124/8mmHg) (right panel). Q-LV: time interval between the beginning of the native QRS complex and the local LV electrogram; LV dP/dt_max_: maximum LV pressure rate

## Discussion

The findings of the present study can be summarized as follows. First, the latest activation site in the LV during RV apical pacing was not the LV posterolateral site but the LV anterolateral site. Second, almost all patients showed that the latest activation site was the basal site in the right anterior oblique view. Finally, simultaneous RV apical and LV anterolateral pacing showed shorter QRSd and better hemodynamic response than simultaneous RV apical and LV posterolateral pacing.

Given that the LV lead position was recommended in the posterolateral to lateral site in a large-scale randomized clinical trial that proved CRT efficacy (1,2), many cardiologists generally place the LV lead at the LV posterolateral site. However, the appropriate LV lead position to restore electrical synchrony depends on the underlying conduction disturbances (3).A recent study showed that in patients with left bundle branch block (LBBB) and left axis deviation, the latest activation site was not the LV posterolateral site but the anterolateral site (11). Therefore, the CRT response was weaker in patients with LBBB and left-axis deviation than in patients with LBBB without left-axis deviation (12,13). This observation implies that the empirical LV lead positioning at the LV posterolateral site was inadequate and that patients with LBBB and left-axis deviation might have more advanced cardiac damage such that the marked conduction delay in the LV is apparent (14), leading to a poor CRT response. The electric LV lead position, assessed by Q-LV or Q-LV ratio, was a predictor of symptomatic and structural responses to CRT in several studies (4,5). Our study showed that the Q-LV and Q-LV ratios were greater at the LV anterolateral site than at the LV posterolateral site during RV apical pacing. Positioning the LV lead at the latest activation site is crucial for improving CRT efficacy. A case of left anterior hemiblock that showed a CRT response was recently reported (15). According to the Q-LV measurement, the LV lead position, in this case, was anterolateral, not posterolateral.

There are many reports on the efficacy of conduction system pacing, such as His-bundle pacing (16) and left-bundle branch pacing (17). However, the RV apical site is still used as the RV pacing site because it is easy for operators to deploy the RV lead and for long-term durability (18). The defibrillator lead is generally placed at the RV apex because of its good defibrillation threshold. When it stimulates the RV apical site, a left-axis deviation always appears. Our study demonstrated that the most recent activation site in patients with RV apical pacing was the LV anterolateral site, as well as in patients with LBBB and left-axis deviation. Our study confirmed that simultaneous RV apical and LV anterolateral pacing showed a narrower QRSd and better LV dP/dt_max_ than RV apical and LV posterolateral pacing. Accordingly, the LV anterior site is highly recommended for LV lead placement in patients who require an upgrade to CRT.

Previous studies that compared biventricular pacing to RV apical pacing have discrepancies. Although the superiority of biventricular pacing was confirmed in the BLOCK-HF trial (19), the results were inconclusive in the BioPace study (20),. Further, there was no statistically significant difference between implantable cardioverter defibrillator and CRT defibrillator among patients who required pacing at baseline in the sub-analysis of the RAFT trial (21). In these three trials, the LV lead position was first recommended in the posterolateral to lateral site whenever possible, and LV lead positioning was modified based on the patient’s CS anatomy. Although the final LV lead position distribution has not been reported, it is assumed that the failure to prioritize the LV anterolateral site for LV lead placement may have caused discordant results.

## Clinical implications

As mentioned above, the LV anterolateral site is preferable for LV lead placement in patients planning to upgrade to CRT. Gold et al. (5) reported an LV lead placement at which a Q-LV over 95 ms was recommended to achieve a good CRT response. In our study, the Q-LV interval at the LV anterolateral site was over 95 ms in all patients; in contrast, the Q-LV interval at the LV posterolateral site was over 95 ms in 30 (81.1%) patients (p=0.01). Duckett et al. (22) reported a significant relationship between an LV dP/dt_max_ increase of >10% in acute hemodynamic studies and LV reverse remodeling 6 months after CRT. In our study, 21 (56.8%) patients showed an LV dP/dt_max_ increase of >10% during simultaneous RV and LV anterolateral pacing, whereas only 10 (27.0%) patients showed an LV dP/dt_max_ increase of >10% during simultaneous RV and LV posterolateral pacing (p=0.02).

Recently, fusion-optimized AV interval programming using residual intrinsic conduction with ventricular pacing has been reported to show a narrower QRSd and better CRT response (23). However, this method does not apply to patients with a long PR interval or an advanced AV block. Patients not eligible for fusion-optimized interval programming should be categorized into the ventricular pacing-dependent group. In this group, the LV anterolateral site is also the preferred LV lead position when the RV is stimulated from the RV apex. Moreover, if conduction disturbances worsen in patients using a fusion-optimized A-V interval and CRT becomes ineffective, LV lead repositioning from the LV posterolateral to the anterolateral site might be considered one of the options to improve the CRT effect.

## Limitations

This study has some limitations. First, the registered patients did not have indications for pacemaker implantation, and patients with underlying conditions such as ischemic heart disease or peripheral artery disease were excluded. However, pacemakers are primarily indicated due to age (24,25), and our registered patients were limited to those aged 65 years or older (average age, 75±8 years). It is difficult to predict which patients will require an upgrade to CRT after pacemaker implantation, but elderly patients who require pacemaker implantation due to right ventricular pacing-induced cardiac dyssynchrony may have the potential to become candidates for CRT upgrade. Although there may be differences in the patient backgrounds between those who meet the criteria for CRT indication and our study population, we believe that our study results should be considered when deciding on the placement of the left ventricular lead during the upgrade procedure. Second, nearly 30% of the patients in this study did not have LV anterolateral or posterolateral branches eligible for LV lead implantation. When there is only one large branch in the LV lateral region, it is inevitable to place the LV lead in the same site. However, this study highlights a new perspective that attention should be paid to the LV anterolateral site. Even if there is only one branch in the LV lateral region, inserting the tip of the LV lead into further branching towards the LV anterolateral direction may be useful. Third, the threshold was too high to stimulate the LV at the latest activation site in 16 patients. In these patients, the pacing site was then moved to a more apical site to achieve sufficient myocardial capture. However, there was no significant difference in Q-LV between the LV anterolateral and posterolateral sites (p=0.85). Fourth, conduction system pacing is an alternative to RV apical pacing. However, conduction system pacing is complex and usually requires the injection of a contrast medium. Some reports have described complications associated with conduction system pacing (26,27). Therefore, RV apical pacing may be a useful option in clinical practice. Finally, our study was limited by its single-center design and small sample size. Further multicenter randomized studies with a larger number of patients are warranted to confirm these findings.

## Conclusion

The LV lead position is preferably at the LV anterolateral site rather than at the conventional LV posterolateral site in patients who are planned to upgrade to CRT from conventional RV apical pacing.

### What is known

Pacing at the latest electrical activation site is crucial to improve electrical dyssynchrony in cardiac resynchronization therapy (CRT).

Typically, in CRT it is recommended to place the left ventricular (LV) lead from the lateral to posterolateral position of the coronary sinus.

### What the study adds

The latest activation site during right ventricular (RV) apical pacing is the LV anterolateral site and not the LV posterolateral site.

Simultaneous biventricular pacing at the RV apex and LV anterolateral site showed a narrower QRS duration and better hemodynamic response than pacing at the RV apex and LV posterolateral site.

The LV anterolateral site is preferable for LV lead position in patients who plan to undergo an upgrade to CRT from conventional RV apical pacing.

## Data Availability

Data and study materials will be available from the corresponding author and other researchers upon reasonable request.

## Acknowledgments

The authors thank Noboru Kitamura and Takeru Takada for technical assistance and support during the pacing study.

## Abbreviations

CRT: cardiac resynchronization therapy
HF: heart failure
LV: left ventricle
CS: coronary sinus
Q-LV: time interval between the beginning of the native QRS complex and the local
LV: electrogram
QRSd: QRS duration
RV: right ventricular
ECG: electrocardiogram
LV dP/dt_max_: maximum LV pressure rate
AV: atrioventricular
LBBB: left bundle branch block

## Graphic Abstract

The electrical and hemodynamic differences with biventricular pacing from the anterolateral compared to the posterolateral branch of the coronary sinus

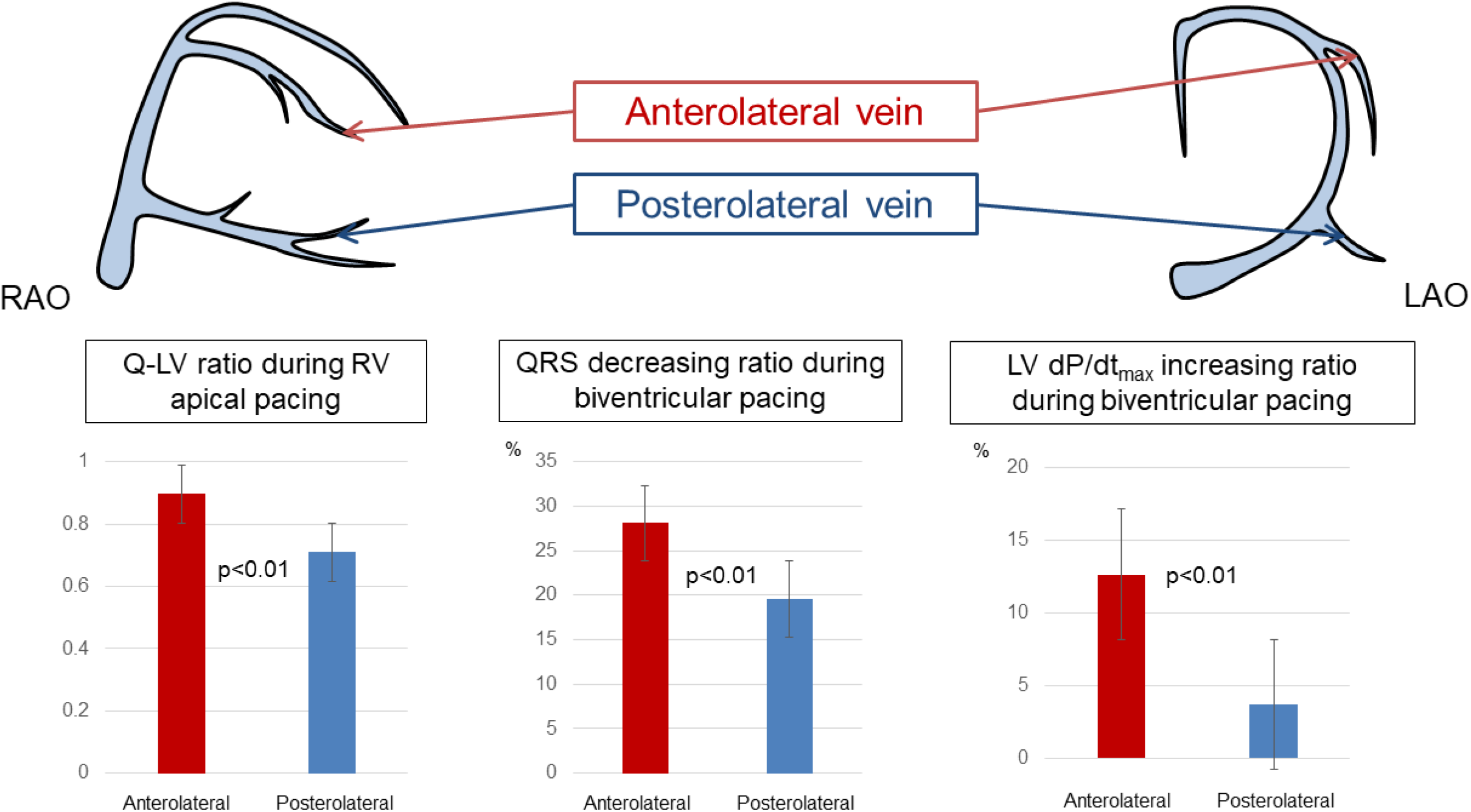
This study demonstrates that the left ventricular (LV) anterolateral site is the preferred position for LV lead placement in patients planned to upgrade to cardiac resynchronization therapy. The Q-LV ratio during right ventricular (RV) apical pacing was longer at the anterolateral LV site than at the posterolateral LV site. The QRS decreasing ratio and LV dP/dt_max_ increasing ratio during biventricular pacing were higher when the LV anterolateral site was used than when the LV posterolateral site was used. Q-LV: time interval between the beginning of the native QRS complex and the local LV electrogram; LV dP/dt_max_: maximum LV pressure rate; RAO: right anterior oblique; LAO: left anterior oblique

